# Trends in medical and surgical admission length of stay by race/ethnicity and socioeconomic status: a time series analysis

**DOI:** 10.1101/2021.05.27.21257925

**Authors:** Arnab K. Ghosh, Mark A. Unruh, Orysya Soroka, Martin Shapiro

## Abstract

Length of stay, a metric of hospital efficiency, differs by race/ethnicity and socioeconomic status (SES). Longer LOS is associated with adverse health outcomes. We assessed differences in average adjusted length of stay (aALOS) over time by race/ethnicity, and SES stratified by discharge destination (home or non-home). Using the 2009-2014 State Inpatient Datasets from three states, we examined trends in aALOS differences by race/ethnicity, and SES (defined first vs fourth quartile of median income by zip code) controlling for patient, disease and hospital characteristics. For those discharged home, racial/ethnic and SES aALOS differences remained stable. Notably, for those discharged to non-home destinations, Black vs White, and low vs high SES aALOS differences increased significantly from 2009 to 2013, more sharply after Q3 2011, the Affordable Care Act’s (ACA) introduction. Further research to understand the ACA’s policy impact on hospital efficiencies, and relationship to racial/ethnic and SES differences in LOS is warranted.

## Introduction

Hospital length of stay (LOS) is used as a management tool to assess both the operational aspects of inpatient treatment and the resources required to deliver care.^1^ There is increasing evidence that hospitals have found the need to actively manage LOS not just for the financial well-being of their systems, but also because of adverse outcomes associated with emergency department boarding,^2^ ambulance diversion due to high bed occupancy,^3^ and delays in discharge.^4^ Furthermore, the Center of Medicare and Medicaid Service’s (CMS) added LOS as a risk-adjusted quality metric to the Physician Value-Based Payment Modifier Program and its successor, the Merit-based Incentive Payment System.^5^

Several studies have revealed racial/ethnic and socioeconomic status (SES) differences in LOS in both medical and surgical admissions, after adjusting for diagnosis, comorbidities, hospital-related factors, and physician-related factors.^6-11^ However, no prior studies have described how these differences vary over time. As hospital systems seek greater market share, higher bed occupancy rates, and increased net revenue,^12^ they may seek efficiencies in LOS to lower costs.^13,14^ Strategies to reduce LOS, including use of multi-disciplinary ward-based activities,^15,16^ changes to work flow management,^17,18^ and designing team structure,^19^ have lowered average LOS from 0.3 to 0.7 days.^16,17,20^ Although the mechanisms driving racial/ethnic and socioeconomic LOS differences remain unclear, it is possible that the strategies implemented may have greater impact on racial and ethnic minorities and patients with low socioeconomic status (SES).

We analyzed an all-payer dataset of hospitalizations from three states for the period 2009 through 2014 to examine differences in adjusted LOS by race/ethnicity and SES over time using robust generalized linear models with fixed effects. Hospitalizations were stratified by discharge destination (home vs non-home destinations) given that patients with higher LOS have a higher likelihood of discharge to non-home destinations. This may be the result of severity of illness, deconditioning as an inpatient, and time required for post-discharge facility placement.^16,21^

## New Contribution

This study adds to the literature on disparities in LOS by race/ethnicity and SES by providing new evidence of how these disparities persist over time using comprehensive inpatient datasets from three diverse states. Furthermore, our rigorous methodological analysis controls for diagnosis, between hospital factors, and patient diagnosis using a novel conceptual framework which outlines the relationship between demographic, hospital, and patient clinical factors and a patient’s LOS. To our knowledge, no other study has utilized these datasets with this new conceptual framework to examine longitudinally whether LOS disparities persist over time.

## Conceptual Framework

Our study employs a novel, evidence-based conceptual framework which details the associations between various factors influencing a patient’s LOS in hospital (Figure 1). These include a patient’s demographic factors (e.g., age, gender, race, insurance status),^8,22,23^ clinical factors (e.g., reason for admission, point of origin of admission [i.e., need for elective vs emergent care], the severity of illness at time of admission, number of comorbidities,^24^ severity of comorbidities, and need for ICU-level care),^25^ social and economic factors (e.g., marital status, income, level of education, adequate housing and support),^9,26-28^ hospital-related factors (e.g., teaching status, for-profit status, hospital size and volume, time of week of admission),^29^ health market dynamics (e.g., bed availability),^30^ and regional practice patterns.^31^

**Figure 1:**
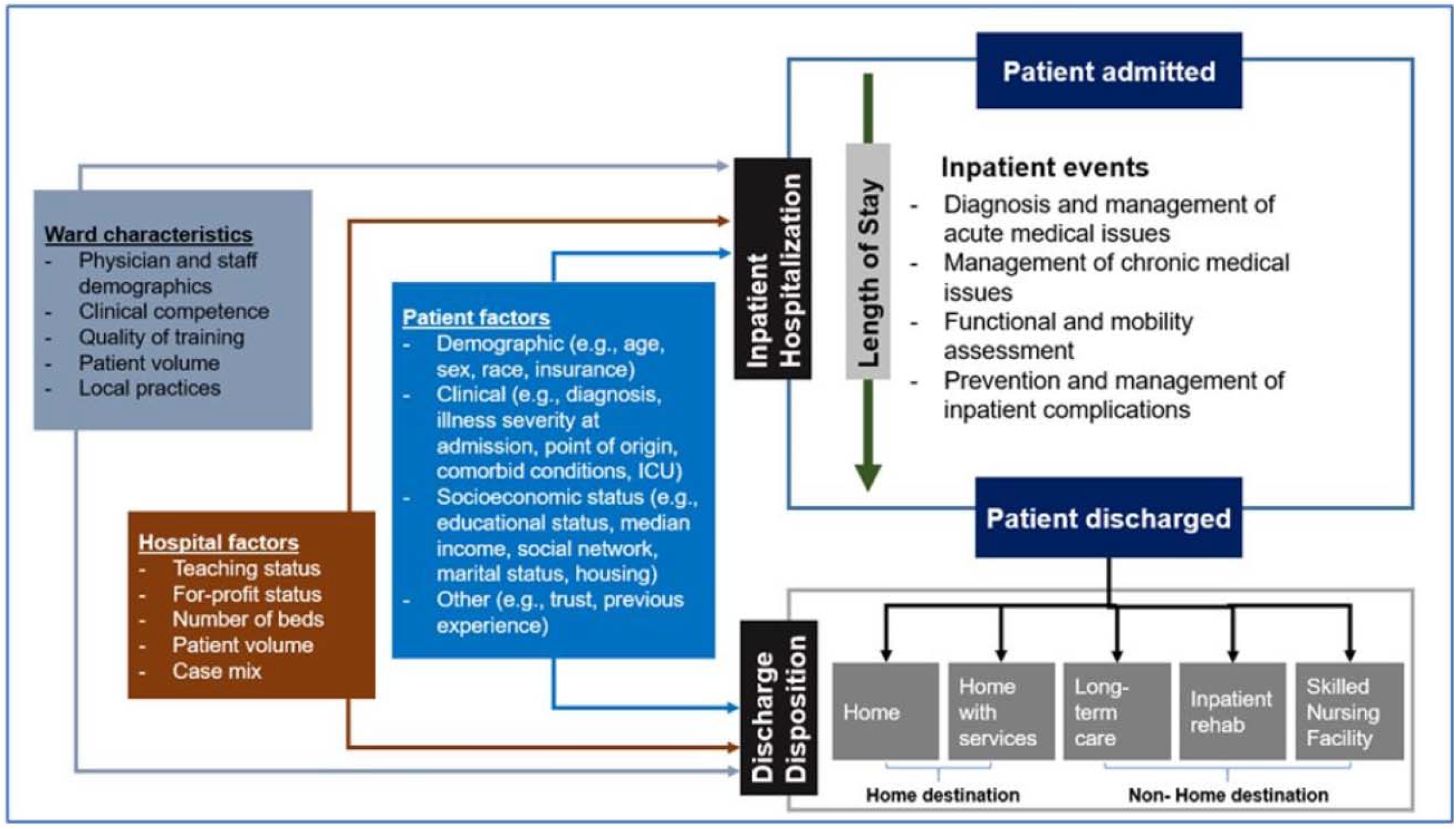
Evidence-based conceptual framework for factors influencing patient length of stay.

Importantly, the patient’s discharge destination plays a prominent role in influencing their LOS. In the United States, patients are typically discharged to one of four locations: home, an inpatient rehabilitation facility, skilled nursing facility (SNF), or long-term care facility. A patient’s discharge destination is a function of a hospital’s ability to improve the health of a patient to the point of discharge; the physician’s opinion that hospitalization is no longer required; and the patient’s clinical and functional needs at the time of discharge, available social supports, and insurance status.

## Methods

### Data Source

We created an analytical file of all-payer inpatient discharges from years 2009 to 2014 using the New York, Florida, and New Jersey State Inpatient Databases from the Health Cost and Utilization project, created by the Agency of Healthcare Research and Quality. This research was approved by the institutional review board of the Weill Cornell Medical College.

### Study Population

The study population consisted of hospitalized patients 18 years of age or older, discharged alive with a medical or surgical diagnosis, based on diagnosis related groups (DRGs) with LOS greater than or equal to 1 day. Although DRGs have been historically categorized by CMS as either medical or surgical, these categories may not reflect the ward where a patient received care. Therefore, we recategorized DRGs by whether they are more likely to be managed on a medical or surgical ward, excluding obstetric, pediatric, and psychiatric diagnoses from the analysis (Supplement). We excluded patients admitted to non-acute care hospitals (long-term acute care hospitals, rehabilitation hospitals, or psychiatric facilities), and discharges from critical access hospitals because of their federally mandated LOS obligations (Supplement).

### Study Outcome

Our outcome was the yearly and yearly-quarter average adjusted LOS (aALOS) by race/ethnicity, and by SES (defined as median income by patient zip code, in quartiles), stratified by discharge destination (home vs non-home destinations [i.e., acute rehabilitation, skilled nursing facilities, long-term acute care hospitals]).

We calculated the aALOS by race/ethnicity and SES over the period, using two separate models informed by our evidence-based conceptual framework (Figure 1). In both models, we treated LOS as a gamma-distributed variable. In the first model which examined racial/ethnic differences, our exposure was race/ethnicity (using White as the reference group). In the second model, our exposure was SES (using low SES [first quartile] as the reference group. In both models, we controlled for patient demographic and socioeconomic characteristics including age, sex, and health insurance (Medicare, Medicaid, private insurance, self-pay), admission-related characteristics (whether admission was on a weekend or not, and whether admission was urgent, elective, or emergent, or other), disease-level characteristics (i.e., number of chronic diseases, Elixhauser-related mortality score), and incorporated separate intercepts for each DRG, each hospital, and each time-quarter to account for differences between DRGs, between hospitals, and seasonality of aALOS. For both models, standard errors were clustered at the level of the hospital (Supplement).

### Statistical Analysis

By exposure (race/ethnicity, and SES), we summarized continuous variables with means and standard deviations or median and interquartile range, where appropriate, and categorical variables with percentages. We assessed differences by race/ethnicity and SES across the range of covariates using ANOVA, Kruskal-Wallis, and chi-square tests where appropriate. Independent variables in both models were checked for multicollinearity using variation inflation factors.

We calculated the aALOS differences by race/ethnicity and SES for each year-quarter of the study period using the *margins* command in STATA on each model. This allowed us to predict the aALOS for admissions overall, and stratified by patients discharged to home, and to non-home destinations based on the previously fit models after averaging out the control variables in the model. To assess racial/ethnic and SES differences in aALOS trends by discharge destination, variance-weighted linear regression was employed. A two-sided α of 0.05 was used to assess statistical significance.

### Sensitivity Analysis

As a sensitivity analysis, the primary analysis was restricted to well-studied inpatient diagnoses of interest to health policy practitioners and CMS: acute coronary syndrome or acute myocardial infarction (ACS/AMI), heart failure, pneumonia, chronic obstructive pulmonary disease (COPD), total knee replacement (TKR), and total hip replacement (THR). The models employed was the same as in the primary analysis, except no individual intercepts for DRGs were used. International Classification of Disease Version 9 codes used for each diagnosis can be found in the Supplement. All analyses were performed in SAS (Version 9.4) and STATA (Version 16).

## Results

### Characteristics of study subjects

In Table 1A and 1B, we summarize the patient characteristics of inpatient admissions to acute care hospitals across the three states from 2009 to 2014 by race/ethnicity (Table 1A), and low vs high SES (Table 1B). In total, we analyzed 22,499,653 admissions. Stratified by racial/ethnic groups (Table 1A), admissions discharged to non-home destinations were, on average, significantly older than admissions discharged home (White: 74.41 vs 62.61 years; Black: 65.08 vs 54.37 years, and; Hispanic: 69.22 vs 56.71 years, all *P* < 0.001), had higher Elixhauser-related mortality scores (White: 7.63 vs 3.64; Black: 7.54 vs 3.41 and; Hispanic: 6.74 vs 2.95, all *P* < 0.001) and more chronic conditions (White: 6.72 vs 5.14; Black: 6.44 vs 4.75 and; Hispanic: 6.05 vs 4.21, all *P* < 0.001), were more likely to be Medicare-insured (White: 79.92% vs 53.08%; Black: 63.62% vs 38.97% and; Hispanic: 68.23% vs 39.49%, all *P* < 0.001), and had a higher unadjusted median LOS (White: 5 vs 3 days Black: 6 vs 3 days and; Hispanic: 5 vs 3 days, all *P* < 0.001)

**Table 1:**
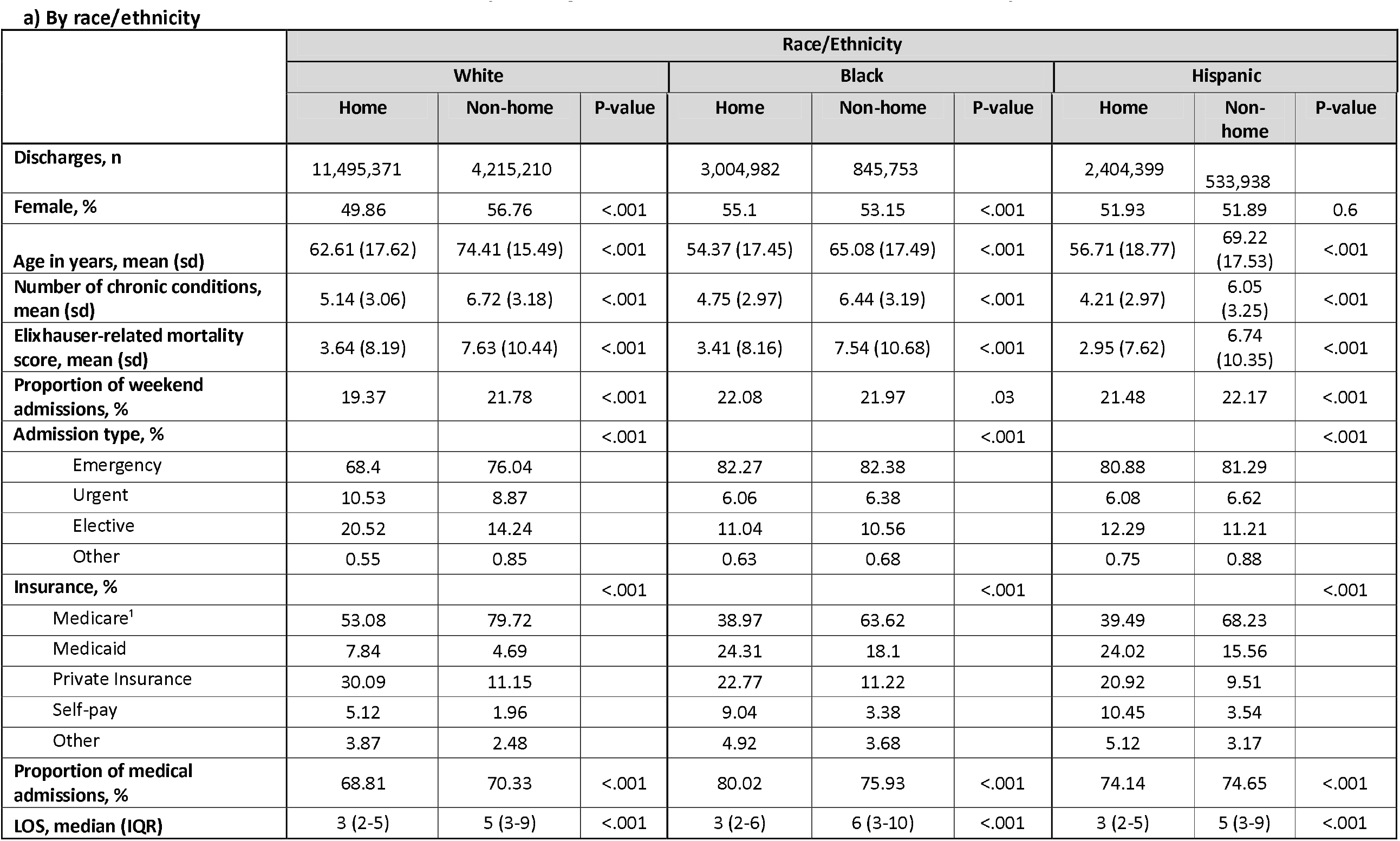
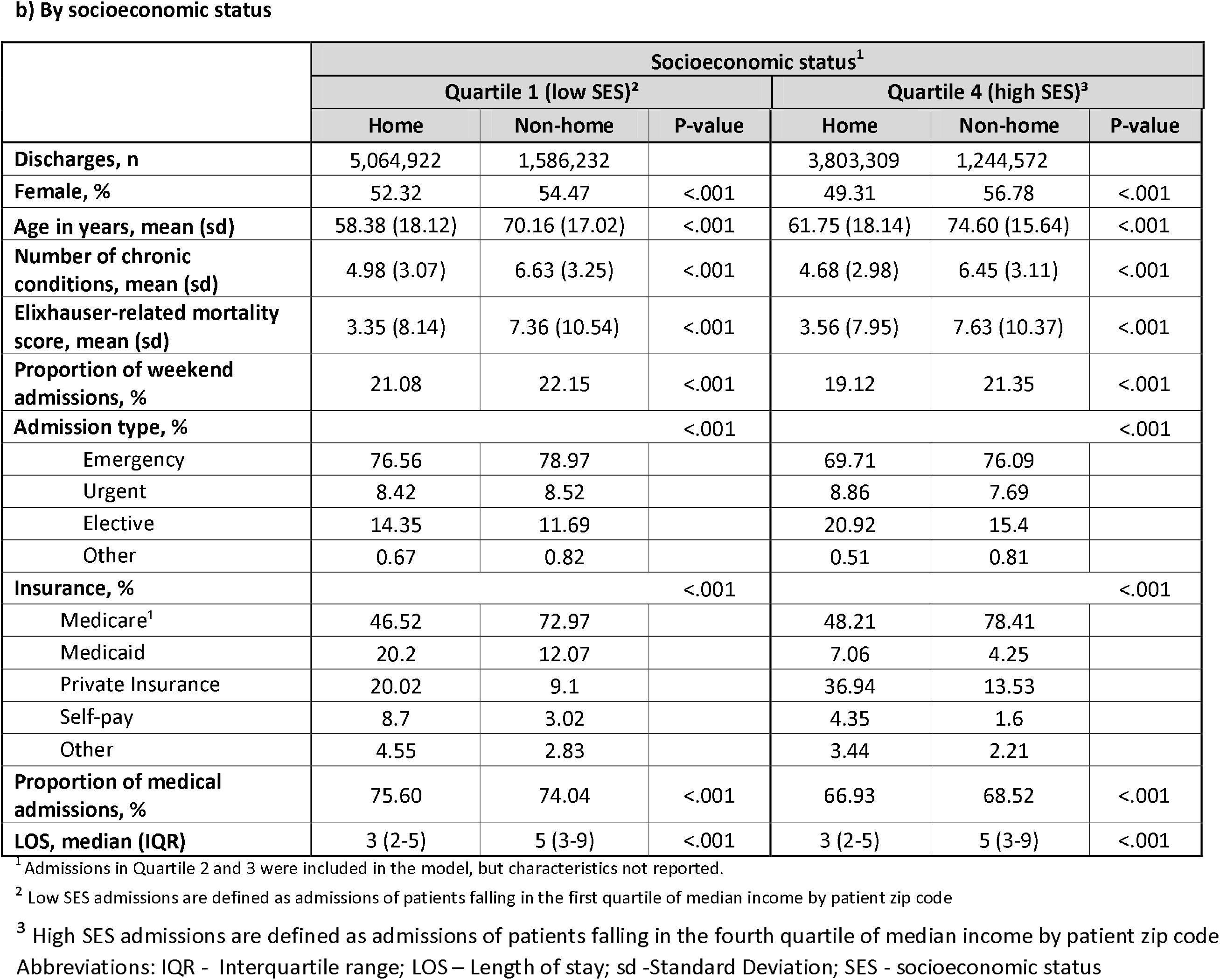
Admission-level characteristics, stratified by discharge destination, New York, Florida, and New Jersey 2009 to 2014

Stratified by SES (Table 1B), admissions discharged to non-home destinations were also, on average, significantly older (low SES: 70.16 vs 53.38 years and; high SES: 74.60 vs 61.75 years, both *P* < 0.001), had more chronic conditions (low SES: 6.63 vs 4.98 and; high SES: 6.45 vs 4.68, both *P* < 0.001), and higher Elixhauser mortality scores (low SES: 7.36 vs 3.35 and; high SES: 7.63 vs 3.56, both *P* < 0.001), were more likely to be Medicare-insured (low SES: 72.97% vs 46.52% years and; high SES: 78.41% vs 48.21%, both *P* < 0.001), and had higher unadjusted median LOS (low SES: 5 vs 3 days; high SES: 5 vs 3 days, both *P* < 0.001).

Table 2 reports the aALOS by year across all racial/ethnic and SES groups by discharge destination. Statistically significant declines were in aALOS were seen for each racial/ethnic and SES group and by discharge destinations (*P-*value for trend < 0.001). The greatest decline from 2009 to 2014 in aALOS occurred among patients discharged to non-home destinations, with high SES patients having an average adjusted decline of 0.79 days (95% CI: -0.80 to -0.79), compared to 0.53 days for low SES patients (95% CI: -0.53 to -0.53). Similarly, LOS for White patients discharged to non-home destinations declined by 0.64 days (95% CI: -0.64 to -0.64), vs. 0.52 days for Hispanics (95% CI: -0.53 to -0.52), and 0.51 days for Blacks (95% CI: -0.51 to -0.51). Declines in aALOS amongst admissions discharged to home decreased by comparable rates across all demographic groups, but by lesser amounts than for non-home destinations. A similar aALOS decline among patients discharged home was found amongst Black and low SES admissions (0.41 day decline, 95% CI: -0.41 to -0.41), Hispanic admissions (0.39 day decline, 95% CI: -0.39 to - 0.39), high SES admissions (0.38 day decline, 95% CI: -0.39 to -0.38) followed by White admissions (0.37 day decline, 95% CI: -0.37 to -0.37).

**Table 2.** Average adjusted length of stay (aALOS)1 from 2009 to 2014 in days, by race/ethnicity and socioeconomic status (SES), stratified by discharge destination2

|  |  | 2009 | 2010 | 2011 | 2012 | 2013 | 2014 | Difference in aALOS, 2009 to 2014 (95% CI) |
| --- | --- | --- | --- | --- | --- | --- | --- | --- |
| **White admissions** |  |  |  |  |  |  |  |  |
| All |  | 5.42 | 5.31 | 5.27 | 5.13 | 5.04 | 4.97 | -0.44 (-0.45 to -0.44) |
| Discharge destination | Home<sup>3</sup> | 4.50 | 4.42 | 4.37 | 4.27 | 4.19 | 4.13 | -0.37 (-0.37 to -0.37) |
|  | Non-Home<sup>4</sup> | 8.13 | 7.96 | 7.93 | 7.69 | 7.57 | 7.50 | -0.64 (-0.64 to -0.64) |
| **Black admissions** |  |  |  |  |  |  |  |  |
| All |  | 5.67 | 5.55 | 5.51 | 5.38 | 5.31 | 5.21 | -0.46 (-0.46 to -0.46) |
| Discharge destination | Home<sup>3</sup> | 4.76 | 4.67 | 4.61 | 4.50 | 4.43 | 4.35 | -0.41 (-0.41 to -0.41) |
|  | Non-Home<sup>4</sup> | 8.28 | 8.05 | 8.08 | 7.93 | 7.85 | 7.77 | -0.51 (-0.51 to -0.51) |
| **Hispanic admissions** |  |  |  |  |  |  |  |  |
| All |  | 5.37 | 5.25 | 5.22 | 5.07 | 4.96 | 4.93 | -0.44 (-0.44 to -0.44) |
| Discharge destination | Home<sup>3</sup> | 4.54 | 4.44 | 4.40 | 4.27 | 4.19 | 4.15 | -0.39 (-0.39 to -0.39) |
|  | Non-Home<sup>4</sup> | 8.00 | 7.83 | 7.88 | 7.65 | 7.49 | 7.47 | -0.52 (-0.52 to -0.52) |
| **Low SES admissions<sup>5</sup>** |  |  |  |  |  |  |  |  |
| All |  | 5.50 | 5.40 | 5.35 | 5.20 | 5.11 | 5.05 | -0.46 (-0.46 to -0.46) |
| Discharge destination | Home<sup>3</sup> | 4.62 | 4.52 | 4.46 | 4.33 | 4.27 | 4.21 | -0.41 (-0.41 to -0.41) |
|  | Non-Home<sup>4</sup> | 8.13 | 7.97 | 7.99 | 7.79 | 7.64 | 7.60 | -0.53 (-0.53 to -0.53) |
| **High SES admissions<sup>6</sup>** |  |  |  |  |  |  |  |  |
| All |  | 5.39 | 5.27 | 5.23 | 5.11 | 5.00 | 4.91 | -0.48 (-0.48 to -0.48) |
| Discharge destination | Home<sup>3</sup> | 4.48 | 4.39 | 4.35 | 4.26 | 4.17 | 4.10 | -0.38 (-0.38 to -0.38) |
|  | Non-Home<sup>4</sup> | 8.16 | 7.95 | 7.88 | 7.65 | 7.52 | 7.36 | -0.79 (-0.79 to -0.79) |

P-value for trend across all admissions by discharge destination was less than 0.001
3 Home defined as discharge to home with or without home services
4 Non-Home defined as discharge to either acute rehabilitation, skilled nursing facilities, long-term acute care hospitals
5 Low SES admissions are defined as patient admissions falling in the first quartile of median income by patient zip code
6 High SES admissions are defined as patient admissions falling in the fourth quartile of median income by patient zip code

Figures 2A to 2C show the trends in racial and SES differences in aALOS by discharge destination by year quarter. Black vs White aALOS differences for discharge home declined very little from 0.27 days in Q1 2009 (95% CI: 0.25 – 0.3) to 0.23 days in Q4 2014 (95% CI 0.21 to 0.26). However, Black-White differences for patients discharged to non-home destinations trended upwards from 0.21 days in Q1 2009 (95% CI: 0.13 to 0.30), reaching its largest difference of 0.34 days in Q3 2013 (95% CI: 0.25 to 0.42). Likewise, low vs high SES aALOS for those discharged home remained stable over the study time period, from 0.14 days in Q1 2009 (95% CI: 0.11 to 0.16) to 0.14 days in Q4 2014 (95% CI: 0.12 to 0.16) but increased for discharges to non-home destinations from 0.03 days in Q1 2009 (−95% CI: -0.04 to 0.10) to 0.26 days in Q4 2014 (95% CI: 0.19 to 0.34). For Hispanic discharges, the picture was different. For those discharged home, aALOS relative to Whites remained virtually identical (0.04 days in Q1 2009, 95% CI: 0.01 to 0.06; 0.05 days in Q4 2014, 95% CI: 0.03 to 0.08). For those discharged to non-home destinations, the relative advantage in terms of shorter aALOS in Q1 2009 (−0.14 days, 95% CI: -0.23 to -0.04) was no longer evident in Q4 2014 (0.01 days, 95% CI: ---0.08 to 0.1).

**Figure 2:**
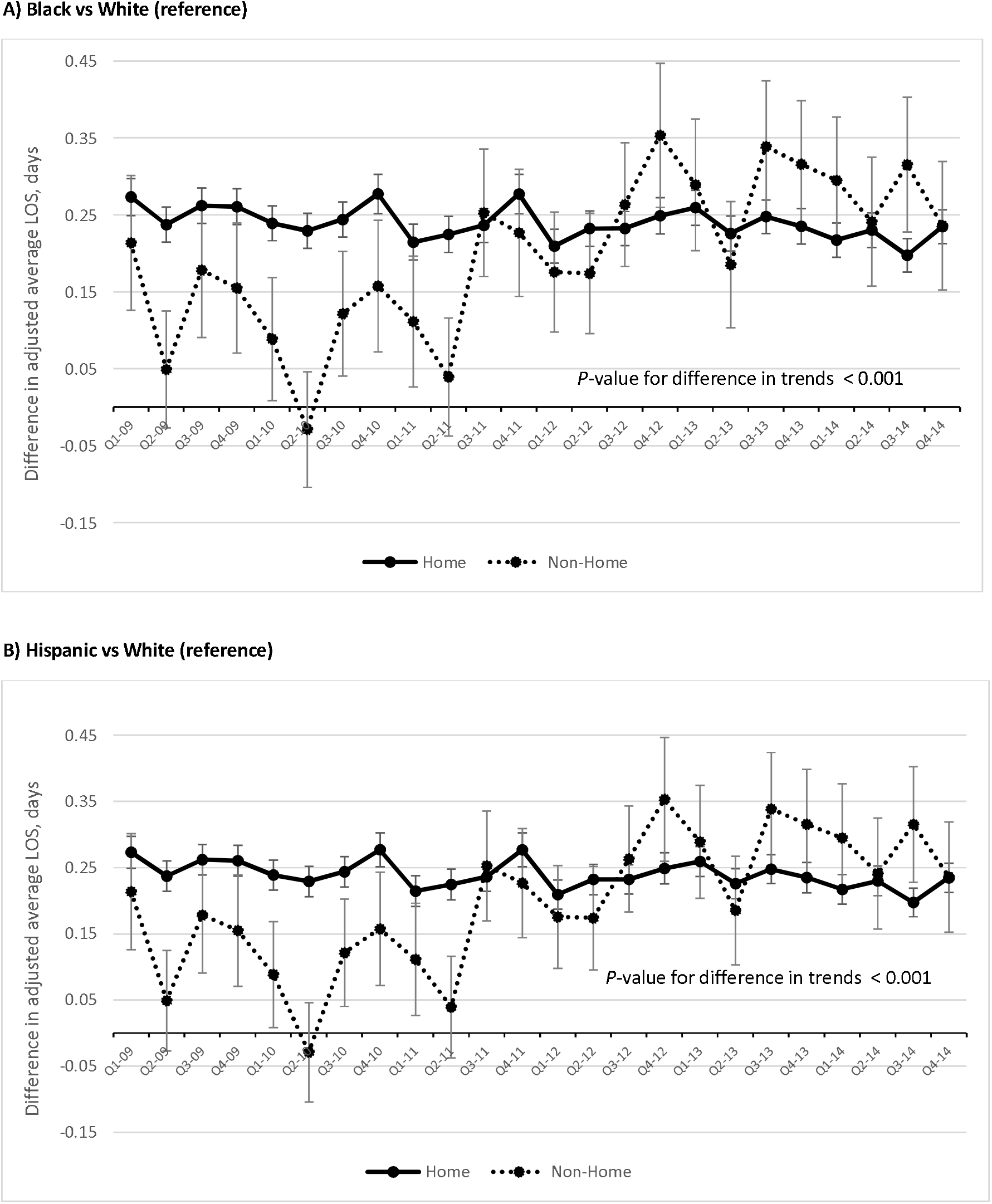
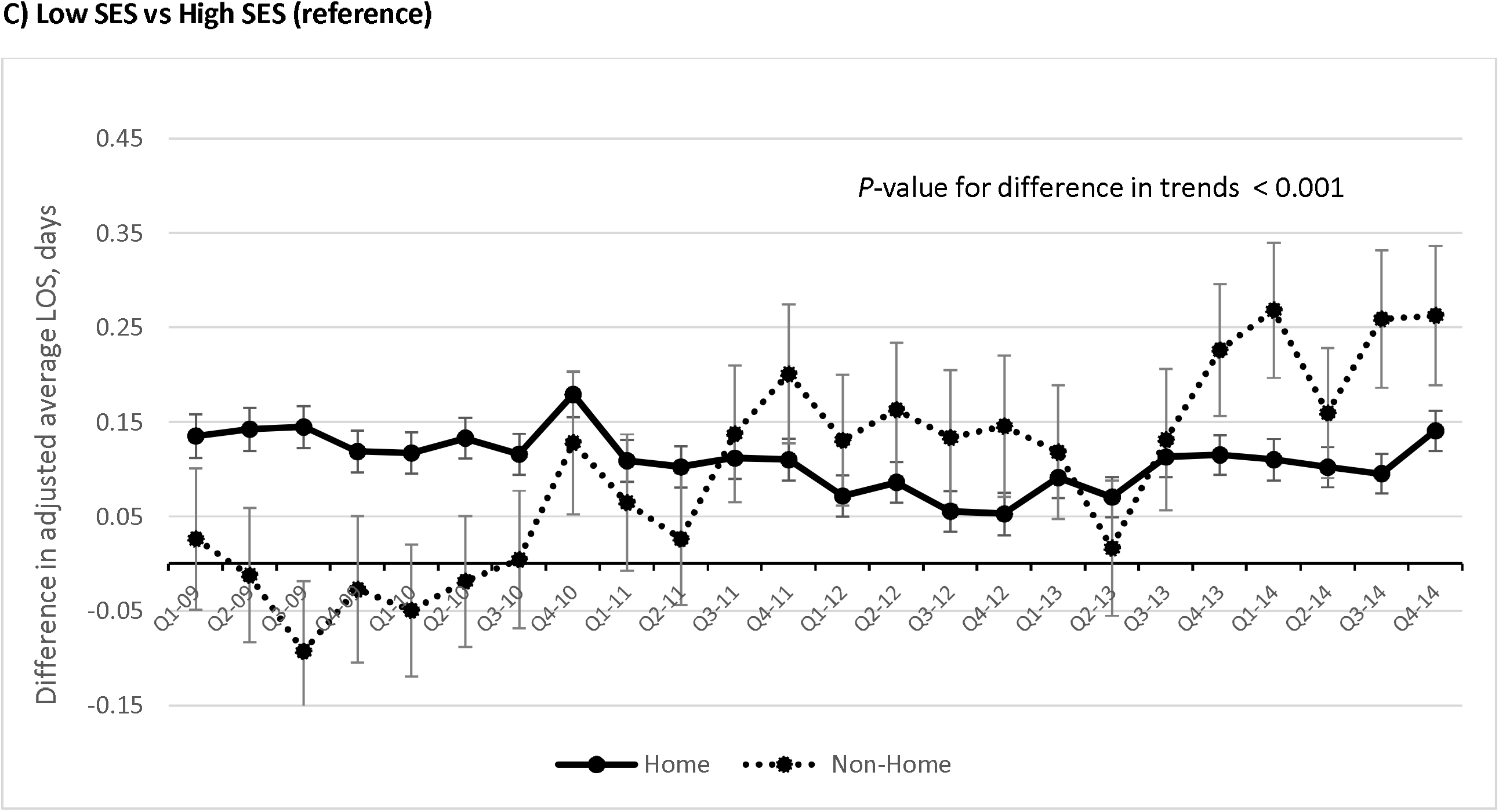
Racial/ethnic and socioeconomic status (SES) differences in average adjusted length of stay (aALOS)^1^ by discharge destination, medical and surgical admissions in New York, Florida, and New Jersey, 2009 to 2014. ^1^aALOS calculated using two multi-variate generalized linear models treating length of stay as a gamma-distributed variable. Model 1 assessed race/ethnicity as the exposure, and controlled for age, sex, health insurance type (Medicare, Medicaid, private insurance, or self-pay), indicator of weekend admission, urgency of admission (elective, emergent, urgent, or other), number of chronic conditions, Elixhauser-related mortality score, and individual intercepts for timequarter, diagnosis-related group, and hospital, with standard errors clustered at hospital level. Model 2 assessed SES as its exposure, with the same control variables as Model 1.

The aALOS trends comparing those discharged to home vs non-home were significant for all three sets of comparisons (*P*-value for trend differences *<* 0.001, Figures 2A-2C). Notably for all three sets of comparisons, compared to the aALOS trend discharged home, the aALOS trend line for discharge to non-home rose and remained elevated after Q3 2011.

### Sensitivity analysis

For individual diagnoses (ACS/AMI, heart failure, pneumonia, COPD, ischemic stroke or transient ischemic attack, TKR or THR), differences in aALOS trends by race/ethnicity and SES did not differ between patients discharged to home and to non-home destinations (Supplement).

## Discussion

In our analysis of an all-payer admission dataset across three states over the period 2009 through 2014, we found that for patients discharged to non-home destinations, trends in racial/ethnic and SES aALOS differences increased significantly. However, they remained stable for those discharged home, and those patients discharged both to home and non-home with well-studied inpatient diagnoses of interest to health policy practitioners and CMS.

To our knowledge, this is the first study to examine trends in adjusted LOS differences by race/ethnicity and SES in a US setting across both medical and surgical admissions. Although evidence is limited to particular patient groups, several studies have highlighted in cross-sectional analyses both racial/ethnic and SES-related differences in adjusted LOS across single institutions, which have demonstrated Black-White differences of 0.4 to 2 days in adjusted LOS in colorectal^8^ and spinal surgery populations,^32^ and SES differences of 0.3 to 0.7 days among trauma patients^22^ and general medical and surgical patients.^33^

Our findings reinforce and generalize the findings from previous studies in three ways. First, our analysis used a large comprehensive dataset of medical and surgical admissions from diverse states in the US which allowed us to detect aALOS differences, compared to studies which have examined racial/ethnic and SES differences within only specific diseases or procedures.^8,9,32,34,35^ Second, because our analysis controlled for differences in admission diagnoses using DRGs our findings have broader applicability to policies focused on LOS metrics at the ward and hospital level, rather than for individuals diagnoses.

Third, neither racial/ethnic nor SES differences were not seen in analyses of trends for individual medical and surgical diagnoses of policy importance. This may reflect the concerted attempts by hospitals (e.g., discharge protocols,^36,37^ checklists, ^37^ care management^38^) to manage quality outcomes, processes and health-related outcomes in these patient subgroups because of related financial penalties. Furthermore, observed racial/ethnic and SES differences in aALOS may reflect the case mix at the ward-level. This is not surprising, given the literature focused on improving patient LOS has focused on large-scale, organizational mechanisms such as patient flow, communication strategies, rather than individual diagnoses, which have little practical applicability on medical and surgical wards which house patients with various conditions.

We found that racial/ethnic and SES differences in aALOS range from 0.15 to 0.25 days for those discharged home, and from 0.10 to 0.35 days for those discharged elsewhere. Given that previously studied strategies to lower average adjusted LOS across wards and healthcare systems led to reductions in average LOS by 0.3 to 0.7 days,^16,17,20^ these persistent differences in aALOS along racial/ethnic socioeconomic lines may be both clinically consequential to patients and financially important for hospitals.

It is unclear why racial/ethnic and SES differences in aALOS remained stable for patients discharged home over the study period but increased for discharges to non-home destinations. The relationship between LOS and discharge to non-home destinations is complex. Patients who spend more time in hospital may be sicker, be subject to more procedures and testing, and thus be subject to more deconditioning. As a result, they may require more intense rehabilitation services to regain sufficient functional improvement before returning to the community. Moreover, discharge to home vs non-home destinations is subject to insurance pre-authorization, patient preferences, and may also depend upon sufficient caregiver support for safe discharge.

Our analysis suggested an inflection point in time where aALOS differences by race/ethnicity and SES begin increasing, starting after Q3 2011. Our study period overlaps with the introduction of the 2011 Affordable Care Act (ACA). As part of its suite of policy changes, several hospital-focused value-based payment models were implemented, including the Center of Medicare and Medicaid Services Hospital Readmission Reduction Program (HRRP). The HRRP sought to penalize hospitals for higher than anticipated readmissions across three targeted conditions, pneumonia, heart failure, and AMI (COPD, coronary artery bypass graft surgery, and elective TKR/THR were added later). Analyses of the outcomes from the HRRP revealed that despite its formal introduction in fiscal year 2013 (October 2012), anticipatory effects related to the changes in hospital adjusted readmission rates began as early as 2011 after the passage of the ACA.^39^ Furthermore, there is evidence to suggest that the effects of the HRRP extend beyond only Medicare Fee-for-service patients to other payer groups,^40^ and non-targeted conditions.^41^ Longer LOS has been associated with lower readmission rates in coronary disease,^42^ in heart failure,^15^ in COPD^43^, and in the general medical and surgical population.^44^ However, other studies in different populations have an inverse association,^45-47^ or an inconsistent relationship for patients discharged to skilled nursing facilities with specific diagnoses.^48^ Whether this policy may be associated with the change in trends of aALOS differences for non-home discharged patients observed in this analysis requires further study.

This study had four limitations. First, our data is limited to only three states. However, the states included in our study have large, diverse populations that should make our findings generalizable. Second, the measure used to denote SES (median income by patient zip code, in quartiles) is only one of many such geographic measures for estimating, and may not reflect its multidimensional nature. Third, the lack of a direct SES measure may also confound the estimated relationship between race/ethnicity and aALOS. Fourth, we did not have information on patient preferences or caregiver support which may influence a patient’s discharge destination, despite controlling for a number of patient, disease, and hospital-related factors.

In conclusion, we found that for hospitalized patients discharged to non-home destinations, racial/ethnic and SES aALOS differences significantly increased between 2009 and 2014 for patients discharged to non-home destinations, and this trend across all three comparisons increased and persisted after Q3 2011. Further research should examine the drivers of increasing differences in aALOS associated with race/ethnicity and SES, including whether hospital-based healthcare policies implemented during the study time period, possibility coinciding with the introduction of ACA, may have been associated with the differences observed in this study.

## Data Availability

Data is publicly available at cost through the Agency for Healthcare Research and Healthcare Quality's Healthcare Cost and Utilization Project

## Acknowledgements

We acknowledge Dr. Benjamin P. Geisler for his assistance in categorizing DRGs.

