## Supplement for "Trends in medical and surgical admission length of stay by race/ethnicity and socioeconomic status: a time series analysis"

**Title**

Arnab K. Ghosh MD, MSc, MA^1^

Mark Unruh PhD, MSc^2^

Orysya Soroka MS^1^

Martin Shapiro MD, PhD, MPH^1^

**Supplement**

eFigure 1. Exclusion Cascade for study

eTable 1: All diagnoses by ICD-9 code

eFigure 2: Racial/ethnic and socioeconomic status (SES) differences in average adjusted length of stay (aALOS) by discharge destination, HEART FAILURE admissions in New York, Florida, and New Jersey, 2009 to 2014

eFigure 3: Racial/ethnic and socioeconomic status (SES) differences in average adjusted length of stay (aALOS) by discharge destination, PNEUMONIA admissions in New York, Florida, and New Jersey, 2009 to 2014

eFigure 4: Racial/ethnic and socioeconomic status (SES) differences in average adjusted length of stay (aALOS) by discharge destination, COPD admissions in New York, Florida, and New Jersey, 2009 to 2014

eFigure 5: Racial/ethnic and socioeconomic status (SES) differences in average adjusted length of stay (aALOS) by discharge destination, ACS/AMI admissions in New York, Florida, and New Jersey, 2009 to 2014

eFigure 6: Racial/ethnic and socioeconomic status (SES) differences in average adjusted length of stay (aALOS) by discharge destination, TKR admissions in New York, Florida, and New Jersey, 2009 to 2014

eFigure 7: Racial/ethnic and socioeconomic status (SES) differences in average adjusted length of stay (aALOS) by discharge destination, THR admissions in New York, Florida, and New Jersey, 2009 to 2014

**eFigure 1. Exclusion Cascade for study**

HCUP hospitalizations in states FL, NJ, NY; years 2009-2014

N = 37,496,626

Age <18 years old

n = 5,308,053

N = 32,188,573

Hospitalizations with patient died

n = 712,323

N = 31,476,250

Hospitalization occurred at CAH, psychiatric, rehab, with RDI=0 hospitals n = 536,236

n=88

N = 30,940,014

Non-acute hospitalizations

n = 5,983,973

N = 24,956,041

Hospitalization’s quarter of year is missing n = 247,572

N = 24,708,469

Length of stay is less or equal 0:

n = 511,107

N = 24,197,362

**eTable 1: All diagnoses by ICD-9 code**

|  | **ICD-9 codes** |
| --- | --- |
| Pneumonia | 480.x, 481.x, 482.x, 483.x, 484.5, 486.x, 487.0, 488.11 |
| Acute coronary syndrome/acute myocardial infarction | 410.xx excluding those with 410.x2 (AMI, subsequent episode of care) 411.1 (unstable angina) |
| Chronic obstructive pulmonary disease | 491.xx - chronic bronchitis, 492.xx - emphysema, 493.2 - chronic obstructive asthma, 496.xx - chronic airway obstruction, not elsewhere classified |
| Heart failure | 402.01, 402.11, 402.91, 404.01, 404.03, 404.11, 404.13, 404.91, 404.93, 428.xx, 398.91 |
| Total knee replacement | 81.54 (procedure code) |
| Total hip replacement | 81.51 (procedure code) |

**eFigure 2: Racial/ethnic and socioeconomic status (SES) differences in average adjusted length of stay (aALOS) ^1^ by discharge destination, HEART FAILURE admissions in New York, Florida, and New Jersey, 2009 to 2014**

A: Black vs White (reference); B: Hispanic vs White (reference); C: Low vs high SES (reference)

**A**

Difference in aALOS, days

|  | Home | Non-Home |
| --- | --- | --- |
| Q1-09 | -0.040589 | -0.299906 |
| Q2-09 | -0.00051 | -0.450045 |
| Q3-09 | -0.075916 | -0.316073 |
| Q4-09 | -0.179582 | -0.37751 |
| Q1-10 | -0.108623 | -0.312338 |
| Q2-10 | -0.101745 | -0.52591 |
| Q3-10 | -0.181399 | -0.25926 |
| Q4-10 | -0.064226 | -0.105323 |
| Q1-11 | -0.214321 | -0.251608 |
| Q2-11 | -0.268902 | -0.377816 |
| Q3-11 | -0.065508 | -0.167282 |
| Q4-11 | 0.000973 | -0.260087 |
| Q1-12 | -0.077311 | -0.381316 |
| Q2-12 | 0.00229 | -0.574908 |
| Q3-12 | 0.049355 | 0.143749 |
| Q4-12 | -0.037133 | -0.07394 |
| Q1-13 | 0.001085 | -0.417939 |
| Q2-13 | -0.040164 | -0.283342 |
| Q3-13 | 0.030804 | 0.311384 |
| Q4-13 | -0.058132 | -0.339885 |
| Q1-14 | -0.05697 | 0.065405 |
| Q2-14 | -0.028472 | -0.358656 |
| Q3-14 | -0.03857 | 0.081881 |
| Q4-14 | 0.011005 | -0.375562 |

Difference in aA LOS, days

|  | Home | Non-Home |
| --- | --- | --- |
| Q1-09 | -0.163702 | -0.40818 |
| Q2-09 | -0.098048 | -0.82806 |
| Q3-09 | -0.103199 | 0.187112 |
| Q4-09 | -0.208188 | 0.094431 |
| Q1-10 | -0.237868 | -0.521202 |
| Q2-10 | -0.061074 | -0.194035 |
| Q3-10 | -0.115926 | -0.516346 |
| Q4-10 | 0.076149 | 0.015344 |
| Q1-11 | -0.181921 | -0.388028 |
| Q2-11 | -0.244674 | -0.399481 |
| Q3-11 | 0.11567 | 0.313461 |
| Q4-11 | -0.096688 | -0.174676 |
| Q1-12 | 0.005823 | -0.362354 |
| Q2-12 | -0.213612 | -0.332124 |
| Q3-12 | -0.067432 | -0.215906 |
| Q4-12 | -0.289705 | 0.142625 |
| Q1-13 | -0.251989 | -0.619143 |
| Q2-13 | -0.076288 | -0.032969 |
| Q3-13 | -0.085691 | -0.265012 |
| Q4-13 | -0.097155 | -0.496473 |
| Q1-14 | -0.125251 | -0.264557 |
| Q2-14 | -0.067751 | -0.573622 |
| Q3-14 | -0.035448 | 0.051984 |
| Q4-14 | -0.073119 | -0.169845 |

Difference in aALOS, days

|  | Home | Non-Home |
| --- | --- | --- |
| Q1-09 | -0.052131 | 0.100625 |
| Q2-09 | -0.027797 | -0.086619 |
| Q3-09 | -0.135744 | 0.059945 |
| Q4-09 | -0.061412 | -0.222466 |
| Q1-10 | -0.016869 | 0.182578 |
| Q2-10 | -0.153643 | 0.305439 |
| Q3-10 | -0.126218 | 0.08653 |
| Q4-10 | 0.111372 | -0.294055 |
| Q1-11 | 0.003889 | -0.10604 |
| Q2-11 | -0.123704 | -0.332657 |
| Q3-11 | 0.040738 | 0.280333 |
| Q4-11 | 0.171044 | 0.145307 |
| Q1-12 | 0.00021 | 0.275488 |
| Q2-12 | -0.043066 | 0.11538 |
| Q3-12 | -0.072446 | 0.214719 |
| Q4-12 | -0.090809 | -0.052729 |
| Q1-13 | -0.123395 | -0.247707 |
| Q2-13 | -0.111078 | 0.104627 |
| Q3-13 | -0.117934 | 0.00786 |
| Q4-13 | 0.02711 | 0.055882 |
| Q1-14 | -0.054864 | 0.184132 |
| Q2-14 | 0.041956 | 0.006188 |
| Q3-14 | 0.027003 | 0.180504 |
| Q4-14 | 0.125753 | 0.100173 |

**eFigure 3: Racial/ethnic and socioeconomic status (SES) differences in average adjusted length of stay (aALOS) ^1^ by discharge destination, PNEUMONIA admissions in New York, Florida, and New Jersey, 2009 to 2014**

A: Black vs White (reference); B: Hispanic vs White (reference); C: Low vs high SES (reference)

**A**

Difference in aALOS, days

|  | Home | Non-Home |
| --- | --- | --- |
| Q1-09 | 0.052808 | 0.052241 |
| Q2-09 | 0.110591 | -0.066699 |
| Q3-09 | 0.208826 | 0.189821 |
| Q4-09 | 0.040875 | 0.011558 |
| Q1-10 | -0.058399 | 0.153257 |
| Q2-10 | 0.117321 | 0.028283 |
| Q3-10 | 0.359694 | 0.413345 |
| Q4-10 | 0.09584 | -0.034893 |
| Q1-11 | 0.093853 | -0.468188 |
| Q2-11 | -0.011884 | -0.027334 |
| Q3-11 | 0.217023 | 0.804366 |
| Q4-11 | 0.232446 | -0.254312 |
| Q1-12 | 0.102909 | -0.016923 |
| Q2-12 | 0.147727 | -0.45361 |
| Q3-12 | 0.182767 | 0.471409 |
| Q4-12 | 0.115819 | 0.111021 |
| Q1-13 | 0.069759 | 0.238951 |
| Q2-13 | 0.082317 | 0.535874 |
| Q3-13 | 0.189283 | -0.041982 |
| Q4-13 | 0.189065 | -0.162035 |
| Q1-14 | 0.01403 | 0.316512 |
| Q2-14 | 0.033138 | 0.235142 |
| Q3-14 | 0.1699 | -0.17314 |
| Q4-14 | 0.167304 | 0.129589 |

Difference in aALOS, days

|  | Home | Non-Home |
| --- | --- | --- |
| Q1-09 | -0.101054 | 0.200671 |
| Q2-09 | -0.034878 | -0.003337 |
| Q3-09 | 0.150218 | 1.16088 |
| Q4-09 | -0.10844 | -0.057411 |
| Q1-10 | -0.06129 | 0.380023 |
| Q2-10 | 0.026315 | 0.038619 |
| Q3-10 | 0.17615 | 0.329209 |
| Q4-10 | -0.136057 | -0.225495 |
| Q1-11 | -0.033972 | -0.195857 |
| Q2-11 | -0.187633 | -0.010975 |
| Q3-11 | -0.03624 | 0.699779 |
| Q4-11 | -0.1143 | 0.164181 |
| Q1-12 | -0.012002 | -0.57286 |
| Q2-12 | -0.071705 | -0.419114 |
| Q3-12 | -0.044275 | 0.079501 |
| Q4-12 | 0.076388 | 0.135865 |
| Q1-13 | -0.208681 | -0.095784 |
| Q2-13 | -0.098431 | 0.515069 |
| Q3-13 | 0.061424 | 0.554366 |
| Q4-13 | -0.036869 | 0.116588 |
| Q1-14 | -0.059563 | 0.057411 |
| Q2-14 | -0.110448 | -0.143515 |
| Q3-14 | -0.047155 | 0.614854 |
| Q4-14 | -0.096401 | -0.245759 |

Difference in aALOS, days

|  | Home | Non-Home |
| --- | --- | --- |
| Q1-09 | -0.151794 | -0.056791 |
| Q2-09 | 0.094221 | -0.014743 |
| Q3-09 | 0.083665 | -0.070698 |
| Q4-09 | 0.061865 | -0.445543 |
| Q1-10 | 0.027982 | -0.284418 |
| Q2-10 | 0.125021 | 0.105018 |
| Q3-10 | 0.191883 | -0.065978 |
| Q4-10 | 0.013351 | -0.257957 |
| Q1-11 | 0.113995 | 0.073799 |
| Q2-11 | 0.184447 | 0.002465 |
| Q3-11 | 0.100648 | -0.117554 |
| Q4-11 | 0.068077 | -0.092306 |
| Q1-12 | 0.105328 | 0.215112 |
| Q2-12 | 0.107588 | 0.01844 |
| Q3-12 | 0.039709 | 0.163633 |
| Q4-12 | -0.009503 | -0.068682 |
| Q1-13 | -0.063766 | -0.138514 |
| Q2-13 | 0.074744 | -0.096645 |
| Q3-13 | -0.001054 | -0.104997 |
| Q4-13 | 0.09578 | -0.025895 |
| Q1-14 | 0.070616 | 0.172733 |
| Q2-14 | -0.003692 | 0.065259 |
| Q3-14 | 0.117119 | -0.045935 |
| Q4-14 | 0.084686 | 0.606255 |

**eFigure 4: Racial/ethnic and socioeconomic status (SES) differences in average adjusted length of stay (aALOS) ^1^ by discharge destination, COPD admissions in New York, Florida, and New Jersey, 2009 to 2014**

A: Black vs White (reference); B: Hispanic vs White (reference); C: Low vs high SES (reference)

**A**

Difference in aALOS, days

|  | Home | Non-Home |
| --- | --- | --- |
| Q1-09 | -0.014272 | -0.343618 |
| Q2-09 | -0.079701 | 0.373587 |
| Q3-09 | -0.047844 | -0.218436 |
| Q4-09 | 0.03451 | -0.572224 |
| Q1-10 | -0.078902 | -0.70216 |
| Q2-10 | -0.091948 | -0.103817 |
| Q3-10 | 0.046563 | -0.237345 |
| Q4-10 | 0.043857 | 0.174944 |
| Q1-11 | -0.047914 | -0.148919 |
| Q2-11 | -0.117617 | -0.3718 |
| Q3-11 | 0.000976 | -0.253981 |
| Q4-11 | -0.063792 | -0.064353 |
| Q1-12 | -0.12902 | -0.090945 |
| Q2-12 | 0.018016 | -0.118408 |
| Q3-12 | 0.138287 | -0.825711 |
| Q4-12 | 0.019505 | -0.269706 |
| Q1-13 | 0.016709 | 0.361099 |
| Q2-13 | -0.018484 | -0.14334 |
| Q3-13 | 0.007563 | -0.139539 |
| Q4-13 | -0.024303 | -0.229728 |
| Q1-14 | 0.050836 | -0.049893 |
| Q2-14 | -0.126905 | -0.614939 |
| Q3-14 | 0.002813 | 0.441215 |
| Q4-14 | 0.003519 | -0.220506 |

Difference in aALOS, days

|  | Home | Non-Home |
| --- | --- | --- |
| Q1-09 | -0.05711 | -0.237724 |
| Q2-09 | 0.029577 | -0.21346 |
| Q3-09 | 0.075465 | 0.414221 |
| Q4-09 | 0.063561 | 0.281177 |
| Q1-10 | -0.10344 | 0.322208 |
| Q2-10 | -0.007006 | -0.267225 |
| Q3-10 | 0.020742 | -0.4538 |
| Q4-10 | -0.079603 | -0.305504 |
| Q1-11 | -0.017086 | -0.252532 |
| Q2-11 | 0.010685 | 0.122789 |
| Q3-11 | 0.13319 | -0.16733 |
| Q4-11 | 0.116064 | -0.055028 |
| Q1-12 | -0.059765 | 0.521043 |
| Q2-12 | -0.063062 | 0.001134 |
| Q3-12 | 0.016665 | 0.238033 |
| Q4-12 | 0.129233 | -0.163702 |
| Q1-13 | -0.082046 | 0.081911 |
| Q2-13 | -0.024868 | -0.030613 |
| Q3-13 | 0.138297 | 0.148471 |
| Q4-13 | -0.161863 | 0.357343 |
| Q1-14 | -0.064866 | -0.384672 |
| Q2-14 | -0.023155 | -0.193351 |
| Q3-14 | -0.002087 | -0.073592 |
| Q4-14 | -0.061733 | 0.033081 |

Difference in aALOS, days

|  | Home | Non-Home |
| --- | --- | --- |
| Q1-09 | 0.006021 | -0.034721 |
| Q2-09 | 0.034518 | 0.058639 |
| Q3-09 | 0.040684 | -0.424129 |
| Q4-09 | 0.048181 | -0.260358 |
| Q1-10 | -0.085693 | -0.147048 |
| Q2-10 | 0.010465 | -0.292985 |
| Q3-10 | -0.039313 | -0.266418 |
| Q4-10 | 0.107737 | -0.128046 |
| Q1-11 | -0.07485 | 0.131794 |
| Q2-11 | -0.211763 | -0.408853 |
| Q3-11 | -0.142561 | 0.075039 |
| Q4-11 | 0.00802 | -0.030638 |
| Q1-12 | -0.022823 | -0.308442 |
| Q2-12 | -0.144797 | 0.104563 |
| Q3-12 | -0.002583 | -0.577183 |
| Q4-12 | -0.049203 | 0.013669 |
| Q1-13 | -0.014248 | 0.133235 |
| Q2-13 | -0.015499 | -0.021907 |
| Q3-13 | 0.14481 | 0.060484 |
| Q4-13 | -0.06 | 0.235174 |
| Q1-14 | -0.066048 | 0.339704 |
| Q2-14 | 0.022313 | -0.06819 |
| Q3-14 | -0.044187 | 0.301444 |
| Q4-14 | 0.050442 | 0.0673 |

**eFigure 5: Racial/ethnic and socioeconomic status (SES) differences in average adjusted length of stay (aALOS) ^1^ by discharge destination, ACS/AMI admissions in New York, Florida, and New Jersey, 2009 to 2014**

A: Black vs White (reference); B: Hispanic vs White (reference); C: Low vs high SES (reference)

Difference in aALOS, days

|  | Home | Non-Home |
| --- | --- | --- |
| Q1-09 | -0.002016 | 0.560685 |
| Q2-09 | -0.046566 | 0.146685 |
| Q3-09 | 0.128256 | 0.915355 |
| Q4-09 | -0.13488 | -0.005476 |
| Q1-10 | -0.041734 | -0.098525 |
| Q2-10 | -0.167749 | 0.235485 |
| Q3-10 | -0.052437 | 0.193747 |
| Q4-10 | 0.001695 | -0.091605 |
| Q1-11 | -0.035513 | -0.23286 |
| Q2-11 | 0.03832 | 0.020594 |
| Q3-11 | 0.1886 | 0.583994 |
| Q4-11 | 0.254714 | 0.579816 |
| Q1-12 | 0.012621 | 0.442056 |
| Q2-12 | 0.121175 | 0.082964 |
| Q3-12 | 0.014946 | 0.584177 |
| Q4-12 | 0.137838 | -0.293036 |
| Q1-13 | 0.067852 | -0.239442 |
| Q2-13 | 0.157775 | 0.078794 |
| Q3-13 | -0.001253 | 0.503315 |
| Q4-13 | 0.077325 | -0.349708 |
| Q1-14 | 0.121749 | 0.443492 |
| Q2-14 | 0.068066 | 0.35525 |
| Q3-14 | -0.107486 | 0.481509 |
| Q4-14 | -0.017363 | 0.27186 |

Difference in aALOS, days

|  | Home | Non-Home |
| --- | --- | --- |
| Q1-09 | 0.16728 | -0.101468 |
| Q2-09 | 0.263055 | 0.266396 |
| Q3-09 | 0.044097 | -0.114898 |
| Q4-09 | -0.06496 | -0.630719 |
| Q1-10 | 0.119598 | -0.222126 |
| Q2-10 | 0.155806 | -0.267252 |
| Q3-10 | 0.102147 | -0.504545 |
| Q4-10 | 0.305478 | -0.702738 |
| Q1-11 | -0.033052 | -0.074754 |
| Q2-11 | 0.045385 | 0.088106 |
| Q3-11 | -0.032964 | 0.156325 |
| Q4-11 | 0.031279 | 0.073084 |
| Q1-12 | 0.206856 | -0.043825 |
| Q2-12 | 0.020293 | 0.262023 |
| Q3-12 | 0.216056 | -0.031365 |
| Q4-12 | -0.028878 | -0.031994 |
| Q1-13 | 0.045729 | -0.284694 |
| Q2-13 | 0.095556 | 0.368059 |
| Q3-13 | 0.117639 | 0.143284 |
| Q4-13 | 0.142839 | 0.105452 |
| Q1-14 | 0.038024 | 0.019925 |
| Q2-14 | 0.241388 | -0.043244 |
| Q3-14 | 0.170252 | -0.115509 |
| Q4-14 | 0.02385 | -0.021933 |

Difference in aALOS, days

|  | Home | Non-Home |
| --- | --- | --- |
| Q1-09 | 0.078014 | -0.173967 |
| Q2-09 | 0.06031 | -0.176289 |
| Q3-09 | 0.131694 | 0.350048 |
| Q4-09 | 0.12474 | -0.218592 |
| Q1-10 | 0.07741 | 0.279386 |
| Q2-10 | -0.060858 | 0.226915 |
| Q3-10 | 0.042862 | -0.316401 |
| Q4-10 | 0.199419 | 0.131503 |
| Q1-11 | 0.008194 | 0.148019 |
| Q2-11 | -0.07552 | -0.12874 |
| Q3-11 | 0.042776 | 0.273367 |
| Q4-11 | 0.052163 | 0.365741 |
| Q1-12 | -0.088147 | 0.132037 |
| Q2-12 | -0.079179 | 0.244367 |
| Q3-12 | 0.172891 | 0.201813 |
| Q4-12 | 0.117615 | 0.218408 |
| Q1-13 | -0.018187 | 0.184435 |
| Q2-13 | 0.077788 | 0.209951 |
| Q3-13 | 0.068769 | 0.289737 |
| Q4-13 | 0.091654 | 0.077945 |
| Q1-14 | 0.005872 | 0.090178 |
| Q2-14 | 0.048316 | 0.466219 |
| Q3-14 | 0.059889 | 0.492291 |
| Q4-14 | 0.075049 | 0.014089 |

**eFigure 6: Racial/ethnic and socioeconomic status (SES) differences in average adjusted length of stay (aALOS) ^1^ by discharge destination, TKR admissions in New York, Florida, and New Jersey, 2009 to 2014**

A: Black vs White (reference); B: Hispanic vs White (reference); C: Low vs high SES (reference)

Difference in aALOS, days

|  | Home | Non-Home |
| --- | --- | --- |
| Q1-09 | 0.329408 | 0.165618 |
| Q2-09 | 0.231824 | 0.179851 |
| Q3-09 | 0.310925 | 0.183226 |
| Q4-09 | 0.29656 | 0.121923 |
| Q1-10 | 0.302167 | 0.151031 |
| Q2-10 | 0.383978 | 0.068712 |
| Q3-10 | 0.30721 | 0.113519 |
| Q4-10 | 0.442257 | 0.124959 |
| Q1-11 | 0.399413 | 0.12951 |
| Q2-11 | 0.243932 | 0.091053 |
| Q3-11 | 0.284722 | 0.101629 |
| Q4-11 | 0.316244 | 0.078276 |
| Q1-12 | 0.279427 | 0.150682 |
| Q2-12 | 0.353047 | 0.164664 |
| Q3-12 | 0.326162 | 0.247597 |
| Q4-12 | 0.186754 | 0.282437 |
| Q1-13 | 0.174795 | 0.104324 |
| Q2-13 | 0.270559 | 0.136216 |
| Q3-13 | 0.194366 | 0.156768 |
| Q4-13 | 0.223951 | 0.103975 |
| Q1-14 | 0.281989 | 0.05287 |
| Q2-14 | 0.189949 | 0.18156 |
| Q3-14 | 0.224355 | 0.007101 |
| Q4-14 | 0.2227 | 0.195939 |

Difference in aALOS, days

|  | Home | Non-Home |
| --- | --- | --- |
| Q1-09 | 0.216449 | 0.175057 |
| Q2-09 | 0.069088 | 0.148676 |
| Q3-09 | 0.216952 | 0.093811 |
| Q4-09 | 0.177807 | -0.057884 |
| Q1-10 | 0.091437 | 0.171984 |
| Q2-10 | 0.332439 | -0.024943 |
| Q3-10 | 0.074516 | 0.109343 |
| Q4-10 | 0.242076 | -0.072994 |
| Q1-11 | 0.153477 | 0.030758 |
| Q2-11 | 0.088026 | -0.024479 |
| Q3-11 | 0.284488 | 0.132199 |
| Q4-11 | 0.236751 | -0.016002 |
| Q1-12 | 0.161508 | 0.078666 |
| Q2-12 | 0.079666 | 0.141204 |
| Q3-12 | 0.043657 | 0.083577 |
| Q4-12 | 0.029097 | 0.050321 |
| Q1-13 | 0.068929 | -0.084526 |
| Q2-13 | -0.005633 | -0.051374 |
| Q3-13 | 0.117822 | 0.046514 |
| Q4-13 | 0.090826 | -0.046697 |
| Q1-14 | 0.046682 | -0.138328 |
| Q2-14 | 0.085949 | -0.033114 |
| Q3-14 | 0.014539 | -0.11896 |
| Q4-14 | 0.135455 | -0.03716 |

Difference in aALOS, days

|  | Home | Non-Home |
| --- | --- | --- |
| Q1-09 | 0.063915 | -0.046525 |
| Q2-09 | 0.023915 | -0.000361 |
| Q3-09 | 0.115245 | -0.02098 |
| Q4-09 | -0.062467 | -0.143043 |
| Q1-10 | 0.004535 | -0.014913 |
| Q2-10 | 0.107146 | -0.032398 |
| Q3-10 | 0.034264 | 0.000781 |
| Q4-10 | 0.095265 | 0.093811 |
| Q1-11 | 0.101564 | 0.134944 |
| Q2-11 | 0.098346 | 0.06157 |
| Q3-11 | 0.105526 | -0.018585 |
| Q4-11 | 0.05055 | 0.038653 |
| Q1-12 | 0.071177 | 0.078762 |
| Q2-12 | 0.046903 | 0.07185 |
| Q3-12 | 0.033272 | 0.101194 |
| Q4-12 | 0.019954 | 0.120935 |
| Q1-13 | 0.005851 | 0.073139 |
| Q2-13 | 0.09736 | 0.082121 |
| Q3-13 | 0.047269 | 0.082142 |
| Q4-13 | 0.100607 | 0.063087 |
| Q1-14 | 0.117912 | 0.043729 |
| Q2-14 | 0.053054 | 0.042826 |
| Q3-14 | 0.037528 | -0.127725 |
| Q4-14 | 0.051229 | 0.032893 |

**eFigure 7: Racial/ethnic and socioeconomic status (SES) differences in average adjusted length of stay (aALOS) ^1^ by discharge destination, THR admissions in New York, Florida, and New Jersey, 2009 to 2014**

A: Black vs White (reference); B: Hispanic vs White (reference); C: Low vs high SES (reference)

Difference in aALOS, days

|  | Home | Non-Home |
| --- | --- | --- |
| Q1-09 | 0.410642 | 0.403462 |
| Q2-09 | 0.496463 | 0.167978 |
| Q3-09 | 0.540641 | 0.517419 |
| Q4-09 | 0.372806 | 0.343468 |
| Q1-10 | 0.599486 | 0.124257 |
| Q2-10 | 0.407015 | 0.341166 |
| Q3-10 | 0.400132 | 0.049306 |
| Q4-10 | 0.661333 | 0.106905 |
| Q1-11 | 0.38665 | 0.152549 |
| Q2-11 | 0.221795 | 0.269082 |
| Q3-11 | 0.123057 | 0.228109 |
| Q4-11 | 0.460451 | 0.093151 |
| Q1-12 | 0.120286 | 0.195561 |
| Q2-12 | 0.423793 | -0.003409 |
| Q3-12 | 0.280499 | 0.273461 |
| Q4-12 | 0.338982 | 0.149584 |
| Q1-13 | 0.345244 | 0.098042 |
| Q2-13 | 0.488735 | 0.033736 |
| Q3-13 | 0.392448 | 0.261798 |
| Q4-13 | 0.195912 | 0.045736 |
| Q1-14 | 0.175131 | 0.183911 |
| Q2-14 | 0.189768 | 0.210919 |
| Q3-14 | 0.231338 | 0.183563 |
| Q4-14 | 0.289246 | 0.093861 |

Difference in aALOS, days

|  | Home | Non-Home |
| --- | --- | --- |
| Q1-09 | 0.238331 | 0.119896 |
| Q2-09 | 0.281153 | 0.043601 |
| Q3-09 | 0.302563 | -0.007518 |
| Q4-09 | 0.226693 | 0.111668 |
| Q1-10 | 0.109693 | -0.043562 |
| Q2-10 | 0.119451 | 0.084605 |
| Q3-10 | 0.30673 | -0.059416 |
| Q4-10 | 0.175729 | 0.056428 |
| Q1-11 | 0.30337 | 0.06135 |
| Q2-11 | 0.345065 | 0.184994 |
| Q3-11 | 0.259266 | -0.057052 |
| Q4-11 | 0.154796 | 0.332639 |
| Q1-12 | 0.095036 | 0.036372 |
| Q2-12 | 0.137625 | 0.012168 |
| Q3-12 | 0.15826 | -0.057662 |
| Q4-12 | 0.048149 | 0.032065 |
| Q1-13 | 0.083556 | -0.135819 |
| Q2-13 | 0.181294 | -0.070094 |
| Q3-13 | 0.168472 | -0.025869 |
| Q4-13 | 0.245186 | 0.008213 |
| Q1-14 | 0.127421 | -0.098734 |
| Q2-14 | 0.21422 | 0.101307 |
| Q3-14 | 0.107734 | 0.031355 |
| Q4-14 | 0.174709 | -0.05422 |

Difference in aALOS, days

|  | Home | Non-Home |
| --- | --- | --- |
| Q1-09 | 0.003099 | 0.074833 |
| Q2-09 | 0.111522 | 0.130851 |
| Q3-09 | 0.094276 | 0.031232 |
| Q4-09 | 0.065035 | 0.054095 |
| Q1-10 | 0.007249 | -0.007785 |
| Q2-10 | 0.096238 | 0.130738 |
| Q3-10 | 0.074602 | -0.08804 |
| Q4-10 | 0.15734 | 0.133035 |
| Q1-11 | 0.193872 | 0.080771 |
| Q2-11 | 0.010045 | 0.073154 |
| Q3-11 | 0.167098 | -0.006542 |
| Q4-11 | 0.157852 | 0.08348 |
| Q1-12 | 0.098655 | 0.131471 |
| Q2-12 | 0.108049 | 0.056788 |
| Q3-12 | 0.055332 | 0.046285 |
| Q4-12 | 0.100906 | 0.115025 |
| Q1-13 | 0.121147 | 0.108519 |
| Q2-13 | 0.144081 | 0.039652 |
| Q3-13 | 0.19819 | 0.100492 |
| Q4-13 | 0.205026 | 0.047707 |
| Q1-14 | 0.118804 | 0.069563 |
| Q2-14 | 0.26473 | 0.098612 |
| Q3-14 | 0.171852 | -0.011687 |
| Q4-14 | 0.16278 | 0.286928 |

^1^ aALOS calculated using two multi-variate generalized linear models treating length of stay as a gamma-distributed variable. Model 1 assessed race/ethnicity as the exposure, and controlled for age, sex, SES, health insurance type (Medicare, Medicaid, private insurance, or self-pay), indicator of weekend admission, number of chronic conditions, Elixhauser-related mortality score, and individual intercepts for time-quarter, diagnosis-related group, and hospital, with standard errors clustered at hospital level. Model 2 assessed SES as its exposure, with the same control variables as Model 1.
